# Standardization of weightage assigned to different segments of the hand X-ray for assessment of bone age by the Greulich Pyle method

**DOI:** 10.1101/2023.06.02.23290917

**Authors:** Chirantap Oza, Anuradha V Khadilkar, Pranay Goel, Tim Aeppli, Shruti Mondkar, Nikhil Shah, Nikhil Lohiya, Hemchand Krishna Prasad, Prashant Patil, Neha Kajale, Vaman Khadilkar, Lars Sävendahl

**Affiliations:** Hirabai Cowasji Jehangir Medical Research Institute, Pune, India; Department of Health Sciences, Savitribai Phule Pune University, Pune, Maharashtra, India; Department of Biology, Indian Institute of Science Education and Research Pune, India; Department of Women’s and Children’s Health, Karolinska Institutet, Astrid Lindgren Children’s Hospital, Stockholm, Sweden; Department of Paediatrics, Cloudnine hospital, Malad, Mumbai, India; Consultant Paediatric Endocrinologist, Division of Growth & Endocrinology, Silver Lining Paediatric Super Speciality Centre for Growth Development & Endocrine Care, Lokmat Square, Nagpur, India; Consultant, Department of Paediatric Endocrinology, Mehta multispeciality hospitals, Chennai, India; Consultant Paediatric Endocrinologist, SRCC NH CHILDREN’S Hospital, Mumbai and Apollo Hospital, Navi Mumbai, India; Senior Paediatric Endocrinologist, Jehangir Hospital, Pune and Bombay Hospital, India

## Abstract

**Objectives:** Bone age (BA) assessment is important in evaluating disorders of growth and puberty; the Greulich and Pyle atlas method (GP) is most used. We aimed to determine the weightage to be attributed by raters to various segments of the hand x-ray, namely, distal end of radius-ulna (RU), carpals, and short bones for rating bone age using the GP atlas method.

**Methods:** 692 deidentified x-rays from a previous study (PUNE-dataset) and 400 from the Radiological Society of North America (RSNA-dataset) were included in the study. Mean of BA assessed by experienced raters was termed reference rating. Linear regression was used to model reference age as function of age ratings of the three segments. The root-mean-square-error (RMSE) of segmental arithmetic mean and weighted mean with respect to reference rating were computed for both datasets.

**Results:** Short bones were assigned the highest weightage. Carpals were assigned higher weightage in pre-pubertal PUNE participants as compared to RSNA, vice-versa in RU segment of post-pubertal participants. The RMSE of weighted mean ratings was significantly lower than for the arithmetic mean in the PUNE dataset.

**Conclusion:** We thus determined weightage to be attributed by raters to segments of the hand x-ray for assessment of bone age by the GP method.

## Introduction

Bone age (BA) assessment is an important tool used in medicine, particularly for disorders of growth and puberty as it correlates better with height velocity, muscle mass, menarche and bone mineral mass than chronological age (CA).[1] For the estimation of bone age, a radiograph of the hand and wrist of the non-dominant hand is assessed by using a traditional manual method or by using an automated method.[2] The manual method involves a comparison of obtained radiograph with radiographs in reference atlases; the two most important publications in this field are by Greulich and Pyle (GP) and by Tanner, Whitehouse and Healy (TW).[3,4]

The GP atlas was created based on radiographs of hands of Caucasian upper middle-class children living in Cleveland, Ohio, United States of America between the years 1931-1942.[5,6]. The GP method is the most popular method among clinicians and radiologists for the assessment of BA; the assessment is relatively quick and easy to learn but reports suggest that there is considerable inter- and intra-observer variability in the ratings obtained.[2] The GP atlas apart from the standard method (i.e., radiograph of the hand and wrist is compared with a standard radiograph from the GP atlas) also suggests a procedure to assess bone age of a hand x-ray film by the rater by adapting the assessment as per his/ her experience/ preference. As per the atlas, after finding a standard X-ray plate that resembles closely the film to be assessed, the rater should make a detailed comparison of individual bones in the following order: distal end of radius and ulna, carpals, metacarpals, and phalanges. Each bone may either be less or more advanced than its counterpart in the standard plate from the atlas. Thus, it is not merely sufficient to assign a single bone age which corresponds most closely to the majority of bony centres visible in it.[3] There may, in some radiographs, be a considerable difference in the bone age of various hand bones, this is possibly because the tubular bones (metacarpals, phalanges, radius, and ulna) mature by processes in the complex growth plates, while the carpals mature by a simpler process of bone apposition. These two mechanisms of growth of the skeleton are governed by different sets of hormones. The tubular bone maturation is more sensitive to sex hormones than the maturation of the carpals.[7] In a recent study, we observed the mean advancement of carpal bone age to be −1.1±0.4 years which may be due to differences in the ethnicity/environment in Pune (in 2015) vs Ohio (1940).[7]

Further, there is no standardization for assigning weights to bones of the wrist and hand, thus, a rater may assign different weights to different bones as per his/ her experience/ preference. This variation is particularly due to the carpal bones as some raters may ignore the carpals and others may assign 50% weight to the carpals during the assessment. Raters using the carpals reduce their weightage at higher maturity but again, there is no standardized method for assigning these weightages.[8] Variation by ethnicity and earlier development of secondary sexual characteristics due to secular trends further complicates the problem.[9,10,11] That BA estimation using the GP method may be imprecise in children of Indian ethnicity has been documented earlier by our group.[12]

Rating each bone individually and assigning separate weightage to each of the 28 bones of the hand and wrist is cumbersome and time consuming. We thus, similar to the study by Carpenter et al, for the purpose of BA assessment by GP method, divided the hand and wrist bones according to the anatomical differences and maturational disparity, into three segments.[13] We divided the whole hand x-ray film into three segments: 1) the distal ends of radius and ulna, 2) the carpals, and 3) the tubular bones i.e., metacarpals and phalanges.

Given that there is no standard weightage attributed to each bone of the wrist and hand film in the GP atlas method and no particular adjustment for racial/ethnic variations and secular trends exists till date, our study aimed to standardize weightage assigned to segments of the hand while using the GP method of BA assessment. Our specific objective was to determine the weightage to be attributed by raters to various segments of the hand X-ray namely, distal end of long bones of the forearm (radius and ulna), carpals and short bones of hand (metacarpals and phalanges) for rating bone age using the GP atlas method.

## Materials and methods

### Subjects

We extracted (from a previous study, children who had height and weight within reference range for age and gender) [14] deidentified left hand radiographs of 692 healthy Indian subjects aged 2–19 years as per their bone age assessment (referred henceforth as the PUNE dataset) [12] and 400 X-rays from the Radiological Society of North America (RSNA) dataset (available as free download from https://www.rsna.org/education/ai-resources-and-training/ai-image-challenge/rsna-pediatric-bone-age-challenge-2017; referred to henceforth as the RSNA dataset, accessed on May 2022).[15] The original study (PUNE) was approved by the institutional ethics committee and all participants gave written informed consent (original approval was dated 7 Aug 2019). Since we used deidentified data, a waiver was granted by the ethics committee (approval dated 9 July 2020). This study was approved by the Ethics Committee of the Indian Institute of Science Education and Research Pune (IHEC/Admin/2021/014).

### Bone Age Measurement

The mean of the BA assessed by the GP method (using whole hand) by raters was considered as reference rating. True rating was defined as the average rating of a large number of expert raters; the root mean square error (RMSE) of the reference rating relative to the true rating was obtained as the standard deviation of the manual ratings divided by the square root of the number of raters.[7] The mean of the rating of the 3 segments of the X-ray is referred to as the arithmetic mean rating.

For GP atlas assessment of the PUNE dataset, three paediatric endocrinologists (HP, PP and NL) who were skilled in assessment of BA (with >10 years of experience) rated the overall (whole hand) BA. Further, an average of rating by two paediatric endocrinologists (VK and CO-experience 25 years and 5 years respectively) who rated the same X-rays in a segmental manner i.e., assigned a GP age to wrist (distal end of radius and ulna), carpals, and hand bones (metacarpals and phalanges) was computed.[13]

Similarly, for the RSNA dataset, the average of rating by multiple raters was considered reference rating (as available with the RSNA dataset) and two expert paediatric endocrinologists (LS and TA-experience around 25 years and 5 years, respectively) rated the X-rays segmentally.

### Pubertal status

As the details on pubertal status were not available, we considered average age at onset of puberty in girls and boys for the PUNE dataset as 10.5 and 11.5 years, respectively.[16,17] The age at onset of puberty for the RSNA dataset was considered 9.5 and 11 for girls and boys, respectively.[18,19]

### Statistical Analysis

Statistical analysis was performed using the SPSS software for Windows (version 26.0, IBM statistics data editor, IBM Corp., Armonk, NY). We assessed the correlation between arithmetic mean rating by the segmental method and reference rating (the average of our multiple manual ratings as the reference rating) using the Pearson correlation coefficient and the Bland–Altman analysis. Linear regression was used to model reference age as a function of the age ratings of the three segments. The regression analysis was run separately for the PUNE and RSNA datasets, girls and boys, and for pubertal status. All three segments were included as covariates for the below-puberty sub-groups in both girls and boys, while carpals were excluded in the over-puberty sub-groups. Intercept terms were excluded if not significant. Robust regression was used throughout to improve fits. Analysis was carried out in Matlab R2022, using the function “fitlm”. We used step-wise regression to find the best fit combination of variables and weightage in Matlab R2022. Accuracy was defined as “the degree to which the information correctly described the phenomena it was designed to measure.” Accuracy was computed using the root mean square error (RMSE) to assess how close the observed data (reference rating) were to values predicted by the model of weighted segmental rating. To compute RMSE, calculate the residual (difference between prediction and truth) for each data point, compute the norm of residual for each data point, compute the mean of residuals and take the square root of that mean. We applied an independent sample t-test to compare the accuracy of the weighted segmental rating with arithmetic mean rating. The mean absolute deviation (i.e., the average distance between each data point and the mean) of segmental arithmetic mean and weighted mean with respect to reference rating was computed. A *p*-value of <0.05 was considered significant.

## Results

### A) Pune dataset

692 de-identified X-rays of Indian children aged 1 to 19 years were included in the study. The sample composition is as shown in table 1. The regression model results to obtain the reference rating using the weighted segmental method of rating are illustrated in table 2A. Short bones had the highest weightage irrespective of gender and pubertal status (>50%). Carpals were given 27% and 36% weightage in pre-pubertal boys and girls respectively. The radius and ulna segment received 21% and 17% weightage in post pubertal boys and girls respectively. We observed high correlation between arithmetic mean and weighted mean of 0.995. The correlation of arithmetic mean and weighted mean with the reference rating is illustrated in figure 1. It was similar for both methods i.e., 0.989 for arithmetic and 0.990 for weighted mean. The mean absolute deviation of segmental arithmetic mean and weighted mean with respect to reference rating was 0.64 and 0.51 years (*p*<0.05). The Bland Altman plots for arithmetic mean rating and the weighted mean by the segmental method and the reference rating are illustrated in figure 2. The SD among the manual raters was 0.49 years (data not shown), thus, SD between reference and true ratings was 0.28 years. For the Pune dataset, the RMSE of weighted mean ratings was significantly lower (*p*<0.05) than for the arithmetic mean (Table 3A).

**Table 1A:** Study population by bone age and gender for the PUNE dataset.

| Age group | Boys | Girls | Total |
| --- | --- | --- | --- |
| 2 | 12 | 3 | 15 |
| 3 | 19 | 27 | 46 |
| 4 | 41 | 30 | 71 |
| 5 | 35 | 19 | 54 |
| 6 | 18 | 27 | 45 |
| 7 | 26 | 14 | 40 |
| 8 | 18 | 20 | 38 |
| 9 | 24 | 28 | 52 |
| 10 | 14 | 10 | 34 |
| 11 | 17 | 16 | 33 |
| 12 | 21 | 27 | 48 |
| 13 | 25 | 9 | 34 |
| 14 | 24 | 18 | 52 |
| 15 | 14 | 9 | 23 |
| 16 | 9 | 28 | 37 |
| 17 | 21 | 15 | 36 |
| 18 | 20 | 26 | 46 |
| 19 | 8 |  | 8 |
| Total | 366 | 326 | 692 |

**Table 1B:**
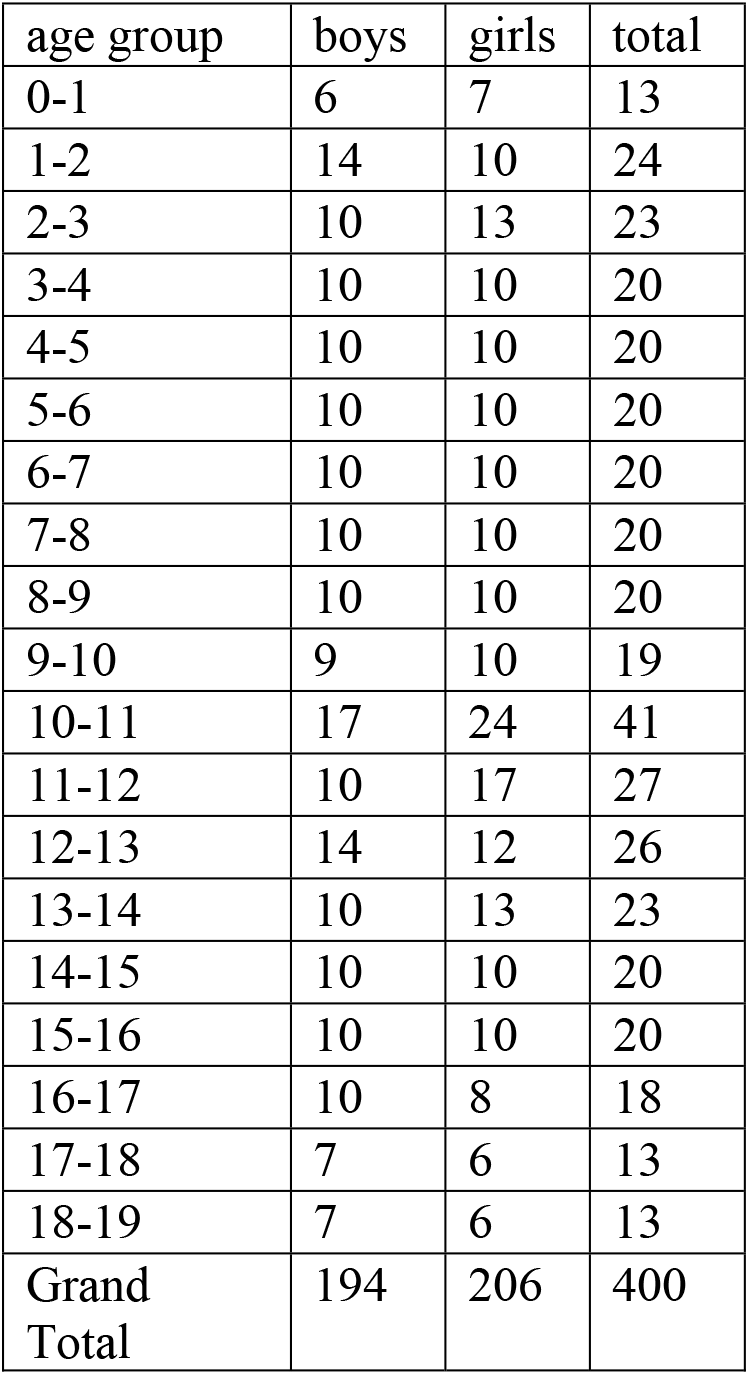
Study population by bone age and gender for the RSNA dataset.

| age group | boys | girls | total |
| --- | --- | --- | --- |
| 0-1 | 6 | 7 | 13 |
| 1-2 | 14 | 10 | 24 |
| 2-3 | 10 | 13 | 23 |
| 3-4 | 10 | 10 | 20 |
| 4-5 | 10 | 10 | 20 |
| 5-6 | 10 | 10 | 20 |
| 6-7 | 10 | 10 | 20 |
| 7-8 | 10 | 10 | 20 |
| 8-9 | 10 | 10 | 20 |
| 9-10 | 9 | 10 | 19 |
| 10-11 | 17 | 24 | 41 |
| 11-12 | 10 | 17 | 27 |
| 12-13 | 14 | 12 | 26 |
| 13-14 | 10 | 13 | 23 |
| 14-15 | 10 | 10 | 20 |
| 15-16 | 10 | 10 | 20 |
| 16-17 | 10 | 8 | 18 |
| 17-18 | 7 | 6 | 13 |
| 18-19 | 7 | 6 | 13 |
| Grand Total | 194 | 206 | 400 |

**Table 2A:** Weightage attributed to segments of wrist and hand films to obtain closest result to reference rating-segregated by gender and pubertal status for the PUNE dataset.

| Age and puberty | Intercept | Radius-ulna | Carpals | Short bones |
| --- | --- | --- | --- | --- |
| Boys<11.5 years | -0.62 | 0.08 | 0.27 | 0.64 |
| Boys>11.5 |  | 0.21 |  | 0.75 |
| Girls<10.5 | -0.45 | 0.11 | 0.36 | 0.51 |
| Girls>10.5 |  | 0.17 |  | 0.81 |

**Table 2B:** Weightage attributed to segments of wrist and hand films to obtain closest result to reference rating - segregated by gender and pubertal status for the RSNA dataset.

| Age and puberty | Intercept | Radius-ulna | Carpals | Short bones |
| --- | --- | --- | --- | --- |
| Boys<11years | 0.16 | 0.23 | 0.15 | 0.56 |
| Boys>11 |  | 0.32 |  | 0.67 |
| Girls<9.5 |  | 0.30 | 0.15 | 0.54 |
| Girls>9.5 |  | 0.30 |  | 0.70 |

**Table 3A:**
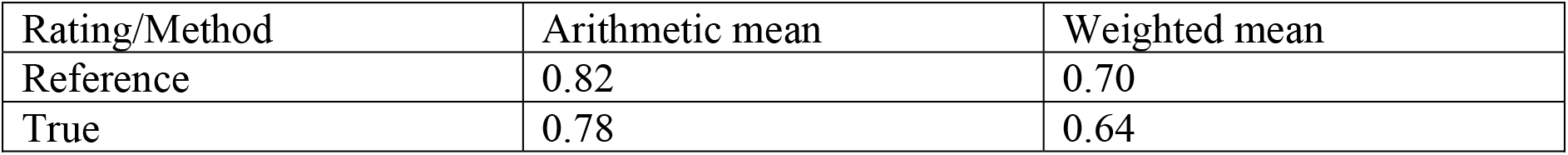
Root mean square errors of the arithmetic mean and weighted mean methods of BA estimation with respect to reference rating and true rating for the PUNE dataset.

### B) RSNA dataset

400 de-identified X-rays of Caucasian children selected randomly (aged 0 to 19 years) were used for the study. The sample composition is as shown in table 1B. The results of the regression model are illustrated in table 2B to obtain reference rating using the weighted segmental rating. Carpals were given 15% weightage in pre-pubertal children irrespective of gender. Short bones received highest weightage in subjects of both genders and pubertal status (>50%). The radius and ulna segment received 32% and 30% weightage in post pubertal boys and girls respectively. We observed very high correlation between arithmetic mean and weighted mean of 0.996. The correlation of arithmetic mean and weighted mean with reference rating is illustrated in figure 3. It was similar for both methods i.e., 0.990 for arithmetic and 0.986 for weighted mean. The mean absolute deviation of arithmetic mean and weighted mean with respect to reference rating was 0.40 and 0.42 years (*p*=0.526). The Bland Altman curves are illustrated in figure 4. The SD among 6 manual raters was 0.68 years, thus, SD between reference and true ratings was 0.31 years.[20] The RMSE were as shown in the table 3B. For the RSNA dataset, there was a trend towards RMSE of weighted mean ratings being higher than that for the arithmetic mean (Table 3B) (p=0.241).

**Table 3B:** Root mean square error of the arithmetic mean and weighted mean methods of BA estimation with respect to reference rating and true rating for the RSNA dataset.

| Rating/Method | Arithmetic mean | Weighted mean |
| --- | --- | --- |
| Reference | 0.70 | 0.84 |
| True | 0.63 | 0.78 |

## Discussion

We found that on regression analysis, carpals were assigned significant weightage in the pre-pubertal age group for both genders in the PUNE and RSNA datasets, however, more weightage to carpals was assigned in the PUNE dataset (27% in boys and 36% in girls) as compared to the RSNA dataset (15% for both genders). The metacarpal and phalangeal segments were assigned the highest weightage in the post-pubertal subjects of both genders in both datasets. In the RSNA dataset, a significantly higher weightage was assigned to the radius-ulna segment (30% for both genders) as compared to the PUNE dataset (21% in boys and 17% in girls). We found a high correlation between weighted mean rating using three segments of the hand and wrist X-ray with reference rating for both the PUNE and RSNA datasets. The RMSE of weighted mean ratings with respect to true rating was significantly lower than for the arithmetic mean for the PUNE dataset while it was higher for the RSNA dataset.

Similar to our study, a study on younger children (less than 10 years old) also reported that in the study group as a whole, hand bone age more closely approximated chronological age than did wrist or carpal bone age. The authors of the study also observed that rapid assessment of bone age using only the carpals or giving higher weightage to carpals lead to under-estimation of the bone age.[13] Moreover, the maturity of carpal bones varies greatly and hence is a major reason for variation in BA estimation amongst raters. We also, in a previous study, observed retardation of carpal bone age as compared to tubular bone age, an effect that was more pronounced in boys.[7] A study to understand maturational disparity between hand-wrist bones in Hong Kong Chinese children by Lee et al., divided the 28 bones of the hand and wrist into six groups so that each group consisted of bones from the same row. The groups were: (1) distal radius and distal ulna, (2) seven carpals, (3) five metacarpals, (4) five proximal phalanges, (5) four middle phalanges, and (6) five distal phalanges. The authors observed that when the mean skeletal age (SA) of all the six groups was considered together, a tendency was observed for the two proximal rows of bones (distal radio-ulna and carpals) to be more retarded, the two distal rows (middle and distal phalanges) were relatively more advanced, and the two groups in-between (metacarpals and proximal phalanges) were intermediate in their maturation. Their SA values of metacarpals and phalanges were very similar to that of the overall SA.[21]

On performing multiple linear regression analysis, we obtained the highest weightage for short bones of the hand i.e., the metacarpals and phalanges. Lee et al. also noted several significant differences between the overall SA and the mean SA of the various bone groups, particularly the carpals. They observed that if the SA of the present group were rated by exclusion of the carpals, a significantly higher estimate of overall SA was obtained, while, if the influence of the carpals was increased by weighing them to as much as 50% of the total score, a considerably lower estimate of the overall SA was obtained.[21] In an attempt to establish backward compatibility to the reference rating using automated methods in children of Indian ethnicity, our group has previously published data on various composite ages with variable weightage on carpal bone age (0, 25, 50, 75 and 100%). We found that the best results were obtained with 50% weightage to carpals. However, these ratings were done using artificial intelligence software and not manually and arbitrary weightage was assigned to carpals and tubular bones.[7] Another study has also observed that a better fit between individual BA and total BA was obtained for distal radius-ulna and metacarpals as opposed to carpals.[22]

A study which aimed to simplify the Tanner–Whitehouse III (TW3) method, formed a grouped-TW algorithm (GTA) wherein Group 1 was composed of the maturity pattern of the radius and the middle phalange of the third and fifth digits and three weights were obtained by data mining, yielding a result similar to that of the TW3 method. Subsequently, new bone-age assessment tables were constructed for boys and girls by linear regression while maintaining accuracy.[23] We used a similar concept but the rating was done using the GP method and weightages as provided by the TW3 method were not used. Also, the previously referred study was conducted on children of Taipei as opposed to our study being performed on children of Indian (Pune dataset) and Caucasian (RSNA dataset) ethnicity. Another Chinese study attributed maturational disparity mainly to middle phalanges and metacarpals. However, they observed that the BAs of the proximal and distal phalanges were closer to the CA in both genders at most ages.[24]

To the best of our knowledge, this is the first study that has established backward compatibility to estimate the weightage attributed to bones of wrist and hands classified in three groups based on anatomy in children of Indian ethnicity. The limitation of this study is lack of details of pubertal status of individual subjects whose X-ray were used in the study, unknown ethnicity of subjects whose data were used from the RSNA dataset and data from a single centre in the Pune dataset that may not be nationally representative. Moreover, validation studies with larger cohorts and multicentric analysis are required to assess the utility of this method of bone age assessment. However, we present a representative case wherein segmental weightage method of BA assessment was used in appendix A.

In conclusion, we have determined the weightage to be attributed by raters to various segments of the hand X-ray for assessment of bone age by the GP method. The weightages differed with ethnicity, gender, and pubertal status. Pediatric endocrinologists and radiologists in India while rating BA by GP method should consider rating the X-rays segmentally and then computing the BA based on weightage assigned.

## Data Availability

All data produced in the present study are available upon reasonable request to the authors.

## Acknowledgement

We thank the Radiological Society of North America for the access to use the imaging datasets and annotations for the purposes of academic research and education, and other non-commercial purposes.

### Appendix A

Example of calculations of bone age by segmental weightage method

**Case 1:**
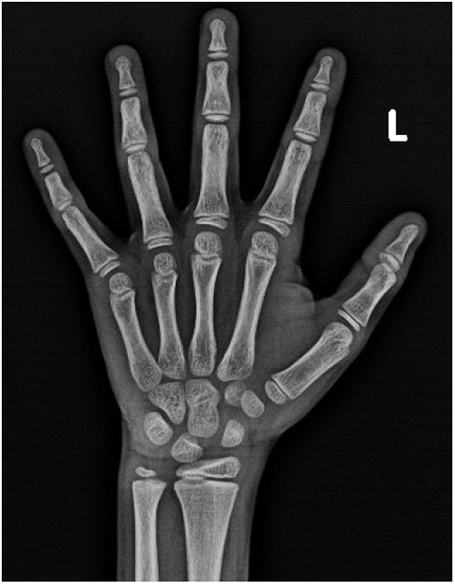
For the x-ray image of boy, the reference rating by GP method was 8.3 years. The GP BA assigned to radius-ulna, carpals and short bones is 8.9, 9.4 and 9.1 respectively. The simple arithmetic mean is 9.1 years while weighted mean is 8.5 years.

---

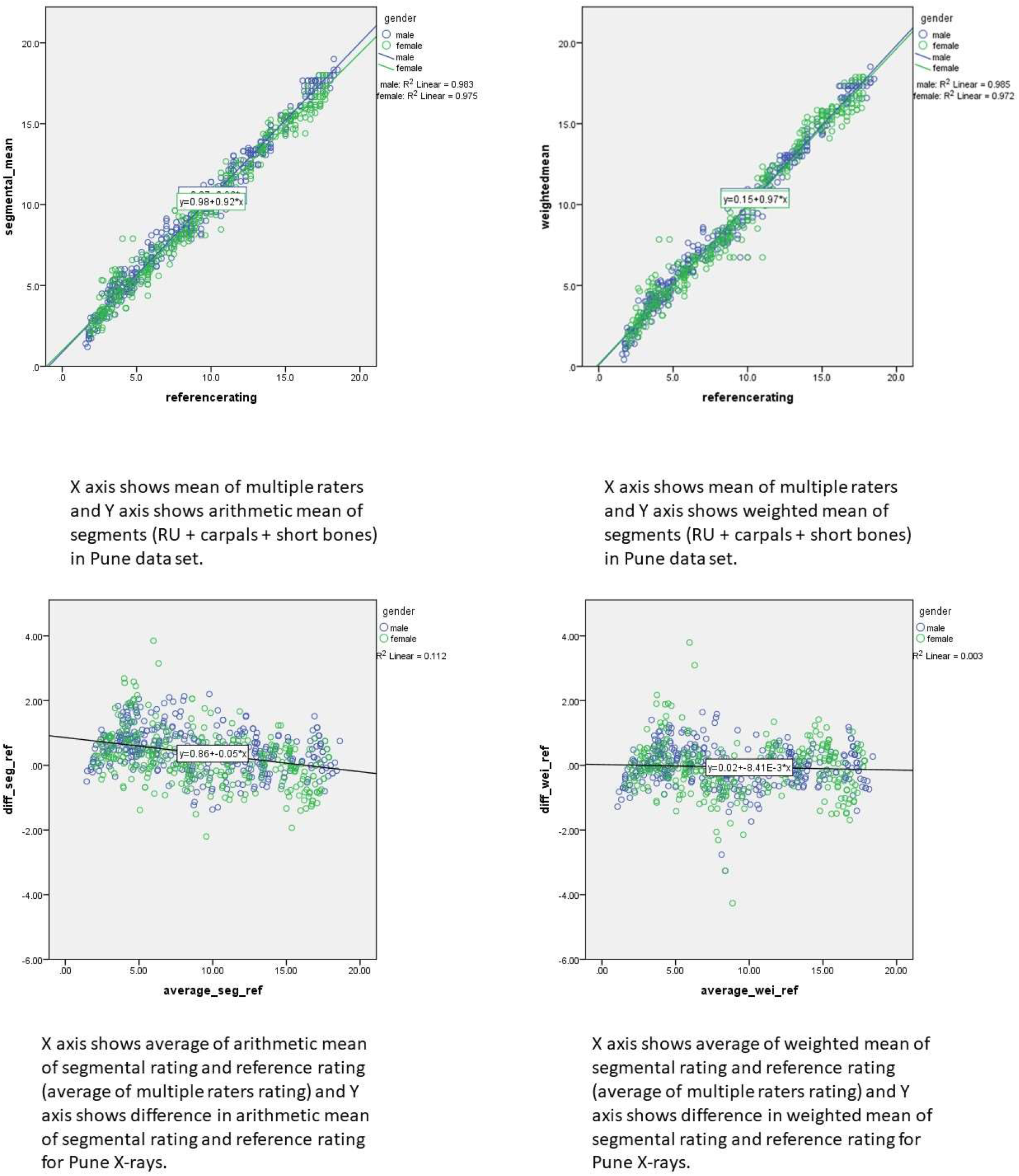

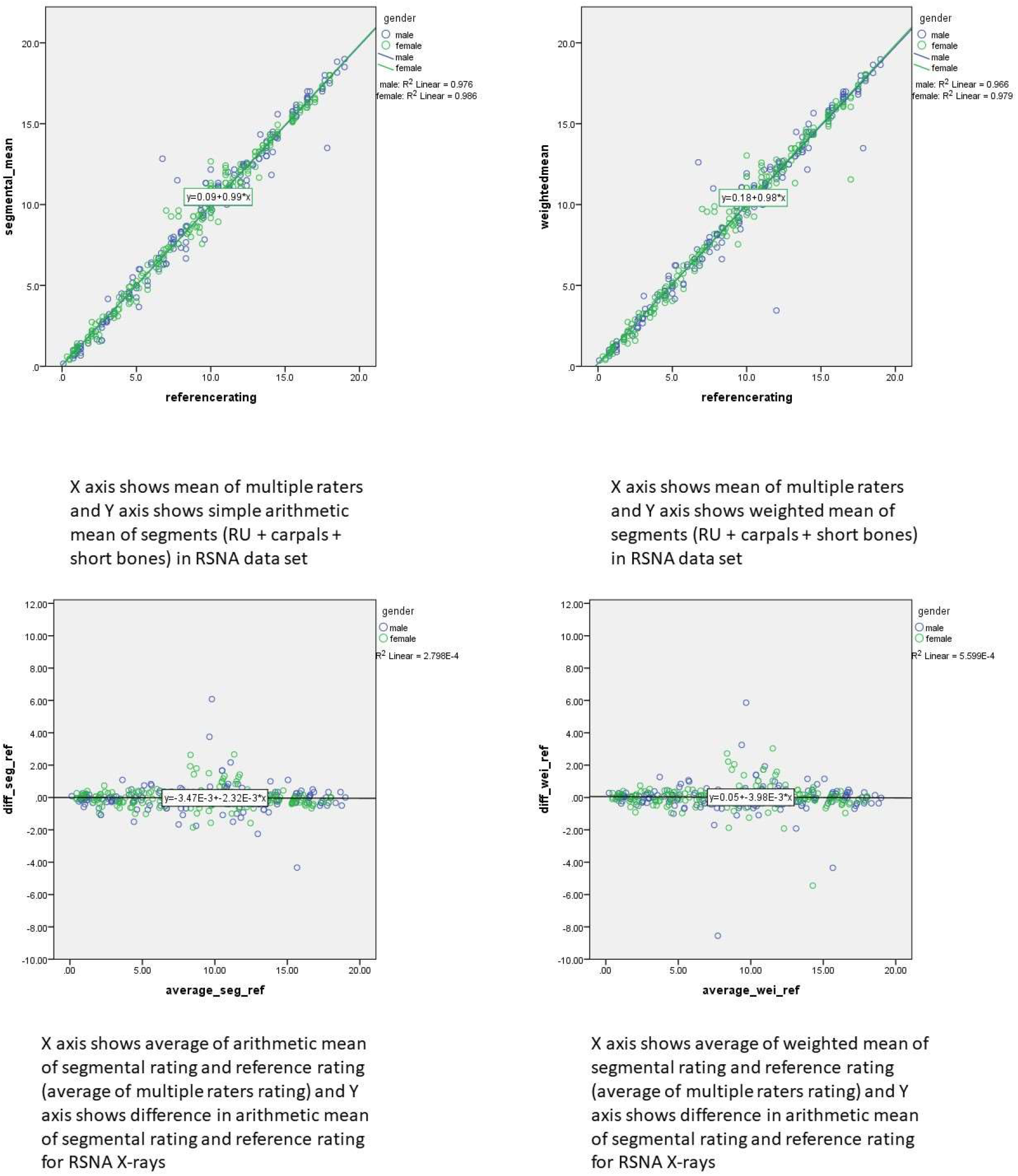

## Notes

### Competing Interest Statement

The authors have declared no competing interest.

### Funding Statement

The project was funded by grants from the Department of Biotechnology (no- BT/PR41073/Swdn/135/4/2020) and Vinnova (no 2020-03609)

### Author Declarations

The original study (PUNE) was approved by the institutional ethics committee and all participants gave written informed consent (original approval was dated 7 Aug 2019). Since we used deidentified data, a waiver was granted by the ethics committee (approval dated 9 July 2020). This study was approved by the Ethics Committee of the Indian Institute of Science Education and Research Pune (IHEC/Admin/2021/014).

